# Prevalence of physical and mental health problems among internally displaced persons in white nile state, sudan 2023

**DOI:** 10.1101/2024.04.16.24305844

**Authors:** Ahmed Mohamed Ahmed Ali, Omran aldoma Mohammed Adam, Sarah Altayeb Mustafa Salem, Sara Hamad Ibrahim Hamad, Nosiba Elnair Musa Ahmed, Hassan Mohamed Ahmed Ali, Aseel Abass Mohamed Abass, Hiba-Allah Abass Mohamed Abass, Abubaker Izz-Eldein Albasher, Mawada Eltagi Elsayid Yousif, Fadwa Elfadil Ahmed Abdalla, Osman AlShazly Osman Abd-alaziz, Samar Bushra Mohammed Ahmed, Shaima Abdelbagi Ahmed Al-Obaid, Abdelgadir H. Osman

## Abstract

**INTRODUCTION:** Despite efforts globally, internally displaced persons (IDPs) face poor living conditions and limited healthcare access compared to refugees. They commonly suffer from malaria, malnutrition, diarrhoea, and respiratory infections, along with mental health issues like depression and PTSD. Severe malnutrition poses significant health risks in the short and long term, especially in developing countries.

**METHODS:** To assess IDPs’ health, we conducted interviews. Out of 574 participants aged 10 and above, many reported physical and mental health problems using questionnaires. Additionally, 226 children under five were surveyed for malnutrition using a questionnaire and measurements of mid-upper arm circumference.

**RESULTS:** The findings revealed that 45% of IDPs experienced poor physical health, with prevalent ailments among adults including minor colds (67%), respiratory infections (43%), prolonged flu episodes (36%), insomnia (38%), headaches (42%), upset stomachs (44%), nausea (35%), and gastrointestinal issues (40%). Depression, PTSD, and anxiety were prevalent among adults at rates of 18%, 20%, and 14% respectively. Moreover, a staggering 72% of children under five suffered from malnutrition, with males accounting for 44% and females 56%.

**CONCLUSION:** In conclusion, the well-being of IDPs residing in camps is detrimentally impacted by the inadequate management, treatment, and prevention of both communicable and non-communicable diseases. Government authorities must recognise and address the complexities associated with multiple agencies involved in delivering healthcare services to IDPs. The implementation of efficient systems to monitor, track, and support IDP healthcare could represent a cost-effective strategy to enhance overall health outcomes.

## INTRODUCTION

Large-scale population displacement often arises from conflicts and natural disasters due to environmental degradation, political persecution, or economic hardships [1]. Internally displaced persons (IDPs) face unique challenges compared to refugees, as they may not receive equivalent assistance from international agencies unless specifically requested by their home country [2].

The consequences of violent conflicts, wars, and displacement events are well-documented, leading to complex humanitarian crises with profound impacts on the well-being of affected populations [3]. These impacts manifest both directly, through violence and physical harm, and indirectly, resulting in increased rates of infectious diseases and malnutrition [4]. Sudan, marked by a history of conflict and instability, experienced another wave of violence in 2023, resulting in a significant number of internally displaced individuals seeking refuge in camps and settlements across the nation [5]. According to the Internal Displacement Monitoring Centre (IDMC), Sudan ranked eighth globally in terms of the number of internally displaced people due to conflict and violence in 2020 [6]. Access to medical care in IDP camps is often limited, leading to preventable deaths and increased morbidity, particularly among women and children [7]. Malnutrition prevails in such settings and significantly elevates the risks of short- and long-term morbidity and mortality [8]. Additionally, conflict-induced displacement increases the risk of mental health disorders, including depression and post-traumatic stress disorder (PTSD), among affected populations [9,10]. The psychological distress stemming from conflicts may also contribute to the adoption of unhealthy behaviours such as hazardous drinking and smoking, leading to a higher burden of non-communicable diseases [11].

The Sudan armed conflict of 2023 resulted in a substantial number of IDPs in the country, many of whom face precarious living conditions and exposure to traumatic events. Despite the evident need, comprehensive data on the prevalence of physical and mental health problems among Sudanese IDPs following the armed conflict are lacking. In order to evaluate and address the health issues facing IDPs, we conducted a cross-sectional study to determine the prevalence of physical and mental health disorders, assess the association between displacement status and mental disorders, and identify the prevalence and associated factors of malnutrition among Sudanese IDPs.

## METHODS

### Participants

The participants in this study were individuals who had been forcibly displaced from their homes due to violent conflict following events in April 2023. They were residing in officially recognised Internally Displaced Persons (IDP) camps in White Nile State. In order to participate, individuals needed to be physically and cognitively capable of completing the questionnaire. Additionally, participants had to be at least 10 years old to take part in the physical and mental health questionnaires, or under 5 years old to participate in the malnutrition questionnaire. Individuals who had been diagnosed with a mental disorder prior to April 15th, 2023 were excluded from the study.

Government statistics indicate that there are over 400 IDPs camps in the state, with around 50,000 inhabitants in total. Approximately 30% of the camp population is under five years old. The state is made up of eight localities, and a sample of 800 IDPs was selected for this study, all of whom agreed to participate in interviews. These participants were from eleven camps located in the four largest localities: Rabak (41%), Kosti (20.5%), Aljabaleen (16.25%), and Aldweem (22.25%). The first two localities contained half of the total camps in the state.

The sample selection method used was stratified random sampling, which involved dividing the population into four localities, selecting the largest camps from each locality, and then randomly selecting participants from each camp to avoid bias. All participants were Muslim and displaced from Khartoum State. Of the 574 participants above ten years old who completed physical and mental health questionnaires, male participants made up 30% of the sample while female participants made up 70%. Among these participants, approximately 48.6% were married, 45.5% were single, 1.4% were divorced, and 4.5% were widowed. Additionally, 226 participants under five completed a malnutrition questionnaire, with male participants representing 45.5% of the sample and female participants representing 54.5%.

Ethical approval was granted by the health research ethical committee of the Ministry of Health, White Nile State before the onset of the study. Participants were approached in their tents and were required to provide informed consent before participating in the study. Confidentiality of information was guaranteed, and all potential subjects were provided with detailed explanations about the study, its purpose, and their rights as participants. This included the right to confidentiality and to withdraw from the study at any time. No names were collected or recorded, and no financial incentives were offered to any participant

### Materials

In 2023, a cross-sectional survey was carried out by the authors and four trained medical student interviewers to gather data from IDPs. A total of 800 interviews were conducted using a face-to-face semi-structured interview method between December 4th and December 14th, 2023. The interviews took place in the camps where the respondents lived, in private outdoor spaces to ensure confidentiality. Each interview lasted approximately 20 minutes on average. This method was chosen to allow respondents to freely discuss sensitive topics without fear of judgment or influence from group dynamics. Unlike group discussions, individual interviews provided a more private and secure environment for participants to express their thoughts and experiences. This approach aimed to capture a more authentic representation of the participants’ perspectives on various issues related to their displacement.

### Participants Above 10

IDPs above 10 responded to three sets of questionnaires -back-translated from English into Arabic- to generate diagnoses of physical and mental health disorders among them. The first questionnaire consisted of 6 questions aimed at gathering demographic information and details about their displacement history. The second questionnaire utilised was the Physical Health Questionnaire (PHQ), a modified version of Spence et al.’s (1987) measure of health. This scale included 14 items that assessed the frequency of symptoms such as sleep disturbances, headaches, respiratory infections, and gastrointestinal problems. Responses were rated on a 7-point frequency scale, with higher scores indicating better somatic health. The total score ranged from 14 to 82, with a mean score of 38.86 (SD 13.32). Participants scoring below 40 were considered to have poor physical health. The third questionnaire used was the MINI (Mini International Neuropsychiatric Interview) was developed by a team of psychiatrists and clinicians led by Dr. Sheehan and Dr. Lecrubier [12]. The MINI is widely recognised and utilised in clinical and research settings for psychiatric assessment and diagnosis. We focused on three modules: Major Depressive Episodes, Post Traumatic Stress Disorder, and Generalised Anxiety Disorder. This comprehensive approach allowed for a thorough assessment of both physical and mental health disorders among the displaced individuals above 10 in the study population.

### Participants under-five

In this study, children and their mothers were recruited to gather information about the children’s nutrition status. The mothers provided oral informed consent for their children to participate in the study, which included a clinical examination for signs of malnutrition such as oedema. The presence of oedema was determined by applying pressure to the feet and checking for a shallow imprint that persisted after the pressure was removed.

The primary method used to assess the nutrition status of children under five years old was a self-administered coded questionnaire that included variables related to dietary intake, general health status, weight for age, weight for height, muscle wasting, water and sanitation access, healthcare access, food security, socioeconomic status, and severity of clinical signs. The GLIM (Global Leadership Initiative on Malnutrition) diagnostic scheme was utilised to assess, diagnose, and grade malnutrition in children. This scheme required at least one phenotypic criterion (such as weight for age or muscle wasting) and one etiologic criterion (such as dietary intake or general health status) to be present for a diagnosis of malnutrition to be made.

In the field of nutrition assessment, measuring malnutrition involves using variables such as weight for height, weight for age, muscle wasting, oedema, dietary intake, and general health status. These variables help evaluate different aspects of an individual’s nutritional status and overall health. Weight for height and weight for age can identify underweight or overweight individuals. Muscle wasting indicates malnutrition and can be assessed through physical examination or imaging techniques. Edema is a sign of severe malnutrition and protein deficiency. Dietary intake is crucial as inadequate consumption can lead to nutrient deficiencies. General health status includes factors like medical history and physical well-being that impact nutritional status. These assessments are considered valid and reliable in clinical practice and research settings.

In this study, we utilised the Statistical Package for Social Sciences (SPSS26) to analyse the quantitative data collected. Descriptive statistics, such as frequencies and percentages, were calculated for all variables in the questionnaire. Chi-squared tests were then employed to explore the relationships between various factors, including physical and mental illnesses, health problems and displacement status, as well as malnutrition and environmental factors. A statistical significance level of p < 0.05 was used to determine the significance of these associations. Furthermore, logistic regression analysis was conducted to assess the risk estimates between depression, PTSD, and anxiety. Crude odds ratios with a 95% confidence interval (CI) were calculated to identify the strength of these relationships. This method allowed us to investigate the potential impact of these mental health conditions on individuals within our study population.

## RESULTS

### Participants above 10

When using a cut-off point of 40 in the physical health questionnaire, our study found that 45% of internally displaced persons (IDPs) were classified as having poor physical health. The most prevalent physical health issues among adults included minor colds (67%), respiratory infections (43%), long-lasting colds or flu (36%), insomnia and disturbed sleep (38%), headaches (42%), upset stomach (44%), nausea (35%), and diarrhoea or constipation (40%). Additionally, the prevalence of depression, PTSD, and anxiety among the participants was 18%, 20%, and 14%, respectively. These findings suggest that IDPs are experiencing a high burden of both physical and mental health issues, highlighting the importance of providing comprehensive healthcare services to this vulnerable population.

In terms of age distribution, the respondents in our study were almost equally divided among three age categories: 31% were aged 11-18 years, 35% were aged 19-35 years, and 34% were above 35 years old. To investigate the relationship between age and physical and mental health problems among internally displaced persons (IDPs), we conducted Chi-square tests. The results showed a significant association between age and poor physical health as well as (PTSD) with a chi-square value of: 12.779, df (2) and P value .002; 9.229, df (2) and P value .010, respectively. This indicates a statistically significant relationship between age and poor physical health and PTSD, at a significance level of p<0.05. However, we found age was insignificantly associated with depression and anxiety among the IDPs.

In terms of educational level, our study found that 11% of respondents had not attended any formal education, while 36.6%, 30.6%, 21.1%, and 1% had education levels of primary, secondary, above educational status, and vocational training, respectively. Upon conducting chi-square tests, the results indicated an insignificant association between educational level and poor physical health, depression, anxiety, and PTSD among the internally displaced persons (IDPs) studied.

We found that male participants accounted for 30% of the sample, while female participants made up 70% of the total participants. The chi-square test results revealed a significant relationship between gender and anxiety, as well as gender and poor physical health, with chi-square values of: 27.695, df (1) and P value .000; 5.888, df (1) and P value.015, respectively which is significant at P<0.05. However, we did not find statistically significant differences between gender and depression or PTSD in this population (p>0.05).

In our study on marital status among IDPs, we found that 48.6% of participants were married, 45.5% were single, 1.4% were divorced, and 4.5% were widowed. Upon conducting statistical analysis using Pearson’s chi-square tests, we discovered a direct relationship between being married and experiencing poor physical health and PTSD among the IDPs. Specifically, the results indicated significant associations between marital status and poor physical health and PTSD, with Pearson’s chi-square value of 12.779, df (2) and P value .002; 9.229, df (2) and P value .010, respectively which is significant at P<0.05. However, we did not observe statistically significant associations between marital status and depression or anxiety in this population.

In our study examining the relationship between displacement period and mental health outcomes among IDPs, we found that the majority of participants, 58%, had been displaced for more than 6 months. A smaller proportion of participants were displaced for 4-6 months (30.1%), 1-3 months (9.6%), and less than one month (2.3%). Interestingly, even though around two-thirds of IDPs experiencing depression, PTSD, anxiety, and poor physical health had been displaced for more than 6 months, our analysis did not detect a statistically significant difference in these health outcomes based on the duration of displacement.

In our investigation into the impact of employment status on the mental and physical well-being of IDPs, we found that the majority of participants (56%) were unemployed. There were also smaller proportions of participants who were students (26.7%), self-employed (5.4%), part-time employees (6.1%), and full-time employees (5.2%). We observed a direct relationship between employment status and poor physical health, depression, PTSD, and anxiety, as indicated by Pearson’s chi-square values. The results showed significant relationships with Pearson’s chi-square values of 19.803, df (4) and P value .001; 19.853, df (4) and P value .001; 10.552, df (4) and P value .032; 10.777, df (4) and P value .029, respectively which is significant at P<0.05.

Figure 1 displays the prevalence rates of physical and mental disorders among IDPs in the White Nile State, providing a visual representation of the burden of these health conditions within the displaced population.

**Figure 1:**
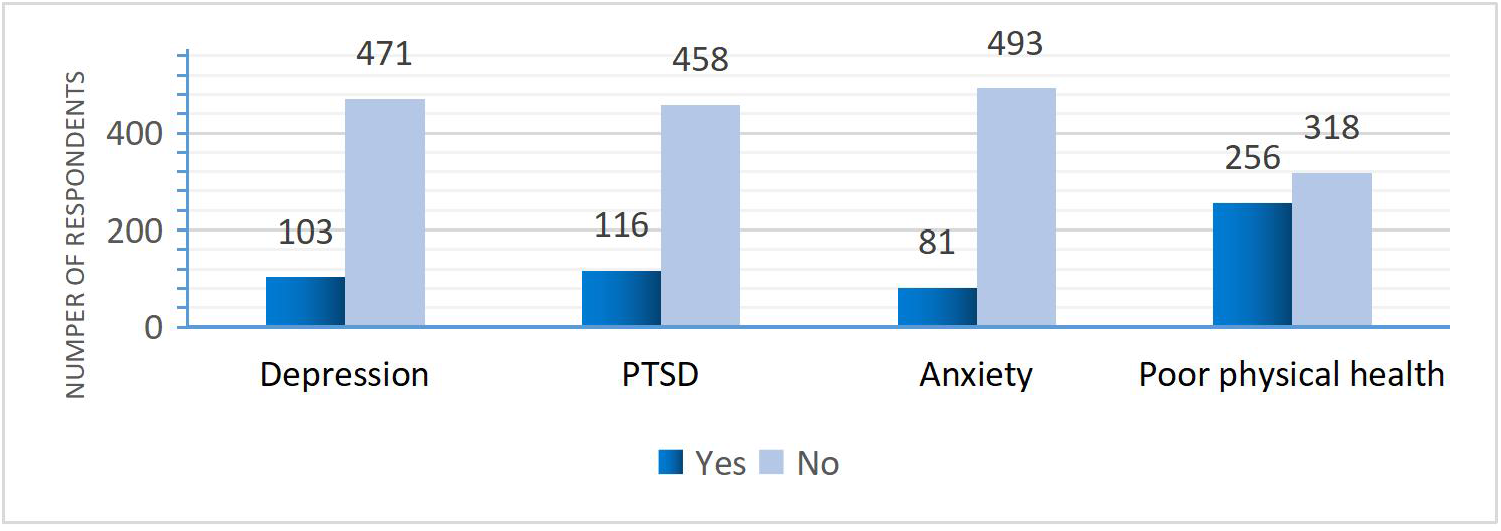
Prevalence of Depression, PTSD, Anxiety, and Poor Physical Health Among IDPs in White Nile displaced camps.

**Figure 2:**
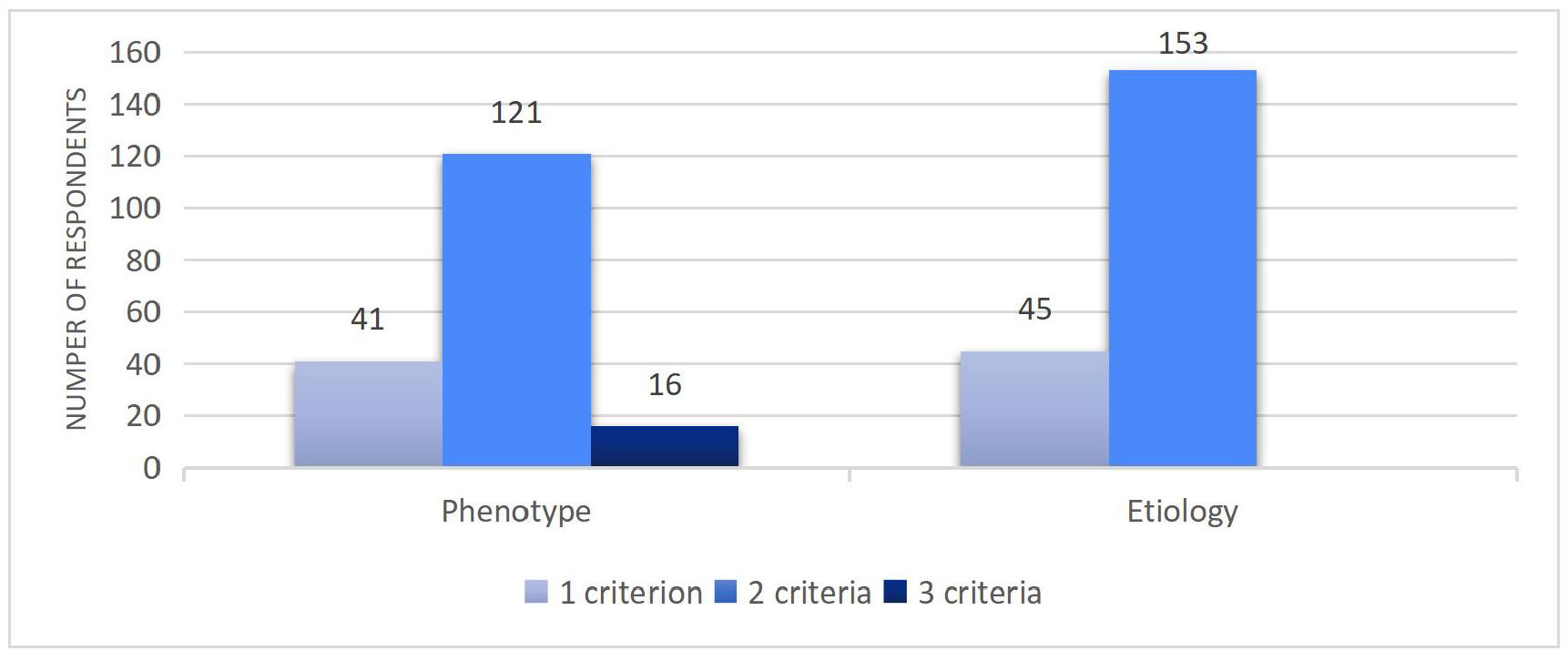
Distribution of phenotypic and etiologic criteria among under-five children in White Nile displaced camps.

The results presented in Table 2 demonstrate a strong correlation between depression, anxiety, PTSD, and poor physical health. The combination of PTSD and poor physical health showed the highest incidence, while the combination of depression and anxiety had the lowest occurrence

**Table 1:**
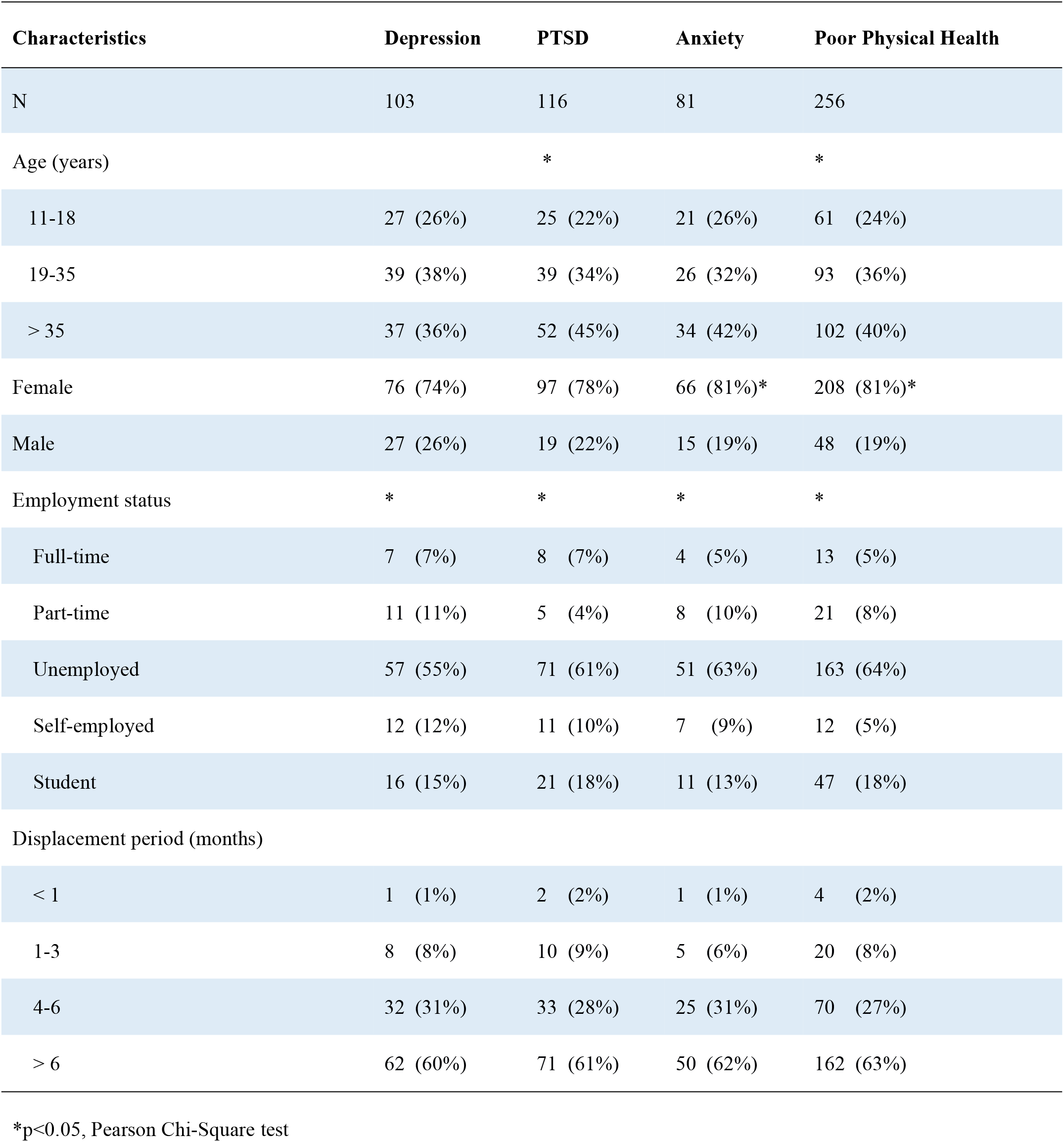
Prevalence of Depression, PTSD, Anxiety, and Poor Physical Health Across Demographic and Employment Characteristics.

| Characteristics | Depression | PTSD | Anxiety | Poor Physical Health |
| --- | --- | --- | --- | --- |
| N | 103 | 116 | 81 | 256 |
| Age (years) |  | * |  | * |
| 11-18 | 27 (26%) | 25 (22%) | 21 (26%) | 61 (24%) |
| 19-35 | 39 (38%) | 39 (34%) | 26 (32%) | 93 (36%) |
| > 35 | 37 (36%) | 52 (45%) | 34 (42%) | 102 (40%) |
| Female | 76 (74%) | 97 (78%) | 66 (81%)* | 208 (81%)* |
| Male | 27 (26%) | 19 (22%) | 15 (19%) | 48 (19%) |
| Employment status | * | * | * | * |
| Full-time | 7 (7%) | 8 (7%) | 4 (5%) | 13 (5%) |
| Part-time | 11 (11%) | 5 (4%) | 8 (10%) | 21 (8%) |
| Unemployed | 57 (55%) | 71 (61%) | 51 (63%) | 163 (64%) |
| Self-employed | 12 (12%) | 11 (10%) | 7 (9%) | 12 (5%) |
| Student | 16 (15%) | 21 (18%) | 11 (13%) | 47 (18%) |
| Displacement period (months) |  |  |  |  |
| < 1 | 1 (1%) | 2 (2%) | 1 (1%) | 4 (2%) |
| 1-3 | 8 (8%) | 10 (9%) | 5 (6%) | 20 (8%) |
| 4-6 | 32 (31%) | 33 (28%) | 25 (31%) | 70 (27%) |
| > 6 | 62 (60%) | 71 (61%) | 50 (62%) | 162 (63%) |

\*p<0.05, Pearson Chi-Square test

**Table 2:** The correlation between depression, PTSD, anxiety, and poor physical health among IDPs in White Nile displaced camp.

| DISORDERS | Depression | Anxiety | PTSD |
| --- | --- | --- | --- |
| Poor Physical Health | 62* | 66* | 83* |
| Depression | - | 25* | 35* |
| Anxiety | - | - | 42* |

\*p<0.05, Pearson Chi-Square test

The Chi-Square test results indicated a significant direct association between depression, PTSD, anxiety with poor physical health. The chi-square values for depression, PTSD, and anxiety were 12.355, 42.742, and 51.921 respectively, all with a P value of .000 which is considered statistically significant at P<0.05. Furthermore, the odds ratios between depression and anxiety, depression and PTSD, as well as PTSD and anxiety were calculated to be 2.37 (95% CI: 1.4%-4%), 2.47 (95% CI: 1.5%-4%), and 6.09 (95% CI: 3.7%-10%) respectively. These findings suggest that individuals with depression, PTSD, or anxiety are more likely to experience poor physical health compared to those without these mental health conditions

## DISCUSSION

Given the rising numbers of internally displaced persons (IDPs) in Sudan and their heightened susceptibility to a range of infectious and chronic diseases, it is crucial to thoroughly document the prevalence of various health issues within these communities. This study represents a pioneering effort in examining the physical and mental health challenges faced by IDPs residing in camps in Sudan. The findings of our study shed light on the intricate relationship between the displacement experience and the health outcomes of IDPs in our study area. Our investigation reveals a concerning prevalence of both physical and mental health issues among this vulnerable population.

Although there have been no recent outbreaks of communicable diseases in the State, we found that a substantial proportion of IDPs exhibited signs of poor physical well-being, with 45% meeting the criteria for classification of poor physical health symptoms. This suggests that the most pressing threats to IDPs are related to their physical safety, human rights violations, and overall well-being. These issues have far-reaching socioeconomic implications for displaced individuals, impacting their health, mental well-being, and ability to reintegrate into society. Among the most commonly reported physical health issues among adults were minor colds, respiratory infections, prolonged cold or flu symptoms, insomnia, headache, upset stomach, nausea, and diarrhoea or constipation. These findings underscore the multifaceted challenges faced by IDPs, ranging from infectious diseases to sleep disturbances and gastrointestinal problems.

Furthermore, our study underscores the prevalence of mental health concerns among IDPs. We found depression impacted 18% of respondents, indicating a substantial burden of depressive symptoms within the population. Additionally, post-traumatic stress disorder (PTSD) and anxiety were reported by 20% and 14% of individuals, respectively. These findings underscore the significant psychological distress experienced by IDPs, likely stemming from the trauma of displacement, loss of home and community, and exposure to ongoing stressors. The observed prevalence of PTSD among IDPs in Sudan, at 20%, is notably higher compared to rates found in similar studies involving Guatemalan refugees (11.2%) and Bosnian refugees (5.6%) [13]. This finding aligns with our initial hypotheses and can be attributed to the severe levels of violence and trauma experienced by the IDPs, including incidents of property destruction, mass killings, air bombardments, and other forms of extreme violence. Additionally, the challenging living conditions within the camps, coupled with high rates of unemployment among the IDPs, likely contributed to the elevated prevalence of PTSD. These results echo previous research findings [14], emphasizing the profound impact of conflict-related trauma on the mental health of displaced populations.

Our analysis delved into the distribution of age among IDPs, revealing intriguing trends across various age groups. Notably, respondents were fairly evenly spread among three age brackets: 11-18 years, 19-35 years, and above 35 years. We found that older age cohorts exhibited a higher propensity to report poor physical health and PTSD compared to their younger counterparts. This observed link suggests that older individuals may encounter challenges in adapting to the new social dynamics brought about by displacement. Older IDPs may struggle to re-establish the social status and roles they held in their previous communities, where responsibilities were clearly defined. This difficulty in adjusting to a changed social environment could contribute to the heightened somatic symptoms and PTSD experienced by older IDPs.

Interestingly, our findings did not show any gender differences in depression and PTSD among IDPs. This lack of distinction may be because both men and women in our study were equally susceptible to mental health issues given the traumatic experiences they had endured. However, previous research suggested that female participants in their study were more vulnerable to developing PTSD due to experiences such as rape, violent loss of family members, and widowhood [15]

To a certain degree, we anticipated similar rates of anxiety in females compared to males based on the relatively similar rates of depression and PTSD between genders. However, our findings revealed that females reported higher levels of anxiety than expected. This suggests that women may be at a heightened risk for developing anxiety disorders, potentially due to a lower threshold for anxiety in females compared to males. This disparity in anxiety levels could be influenced by factors such as the targeted nature of violence against women, specifically in cases of sexual assault. Conversely, the lack of statistically significant differences in depression and PTSD between genders suggests that the manifestation of these conditions may not be solely determined by gender among IDPs. This underscores the complexity of mental health dynamics within displaced communities and calls for comprehensive assessments that consider multifaceted determinants beyond gender alone.

The study underscores a notable association between unemployment and adverse health outcomes among IDPs. We found that a significant proportion of IDPs were unemployed, and this status was linked to higher rates of depression, PTSD, anxiety, and poor physical health. Addressing employment challenges within displaced populations may therefore be crucial in mitigating the burden of mental and physical health issues in such contexts. These findings suggest a significant impact of employment status on the health outcomes of individuals living in camps and heightened vulnerability among the unemployed populace to mental health disorders and physical health issues compared to their employed counterparts.

Although a majority of participants experienced displacement for more than six months, no statistically significant correlation was found between the duration of displacement and the prevalence of depression, PTSD, anxiety, or poor physical health-contrary to common beliefs and previous research findings [16]. This implies that factors beyond displacement duration may exert greater influence on the mental and physical well-being of IDPs. This unexpected result underscores the complexity of factors influencing health outcomes beyond just the length of displacement and challenges the existing notion that longer displacement periods are directly correlated with poorer health outcomes. It is possible that other factors, such as the quality of living conditions, access to healthcare, and social support networks, may play a more influential role in determining the mental and physical well-being of displaced individuals. Further research could explore these non-temporal factors to better understand the complexities of the relationship between displacement period and health outcomes in IDP populations.

Furthermore, our study contributes valuable insights into the nexus between educational attainment, and health outcomes among IDPs. While a substantial proportion of participants had only completed primary education, with a smaller segment having no formal education background, we found no significant correlation between educational level and physical and mental health status. These results suggest that factors beyond education might wield greater influence over mental health outcomes in this vulnerable population.

The discrepancy in the levels of PTSD and somatic symptoms between married and single IDPs contradicts findings from previous research that shows single individuals tend to experience higher levels of psychological distress [17]. Our study reveals a higher prevalence of PTSD and somatic symptoms among married IDPs. This unexpected result may be attributed to the unique challenges faced by married IDPs, who often bear the responsibility of protecting their families amid threats and dangers. The additional pressure on married individuals to ensure the safety and well-being of their loved ones, coupled with difficulties in forming new social networks, could contribute to their heightened levels of distress. While marriage is typically considered a protective factor against psychiatric issues in normal circumstances, the demands of conflict situations or natural disasters may reverse this trend, placing greater burdens on married IDPs [14]. These findings suggest that the context in which individuals live plays a crucial role in determining the impact of marital status on mental health outcomes, highlighting the complex interplay between personal relationships and environmental stressors.

Our study did reveal a strong association between depression, PTSD, and anxiety among IDPs, with individuals experiencing one of these conditions being more likely to also have the others. Specifically, the odds of having PTSD were 2.5 times higher for respondents with depression and 6 times higher for those with anxiety. Additionally, the odds of having depression were 2.4 times higher for individuals with anxiety. This correlation indicates higher odds of experiencing poor physical health in the presence of depression, anxiety, or PTSD and underscores the intertwined nature of mental and physical well-being, with PTSD showing the strongest association with poor physical health among the studied mental health issues. This interconnectedness highlights the complex relationship between these mental health disorders in displaced populations

Turning to the findings concerning children under five years old, malnutrition emerged as the predominant physical health issue in this demographic. The disparities observed in malnutrition rates underscore the multifaceted nature of the problem, with factors such as access to healthcare, food security, and socioeconomic status playing pivotal roles. Notably, the absence of a significant disparity in malnutrition rates between boys and girls in this age group highlights the universal impact of malnutrition across genders.

Our research findings indicate a concerning trend, showing that out of every ten children surveyed, seven were found to have malnutrition. This represents a significant increase in the prevalence of malnutrition compared to previous studies. Despite the potential availability of food, several factors may explain these troubling results, including elevated food prices that exceed normal levels. Research conducted among suddenness refugees has demonstrated a concerning association between the prevalence of communicable diseases such as malaria, diarrhoea, and pneumonia, and an increased risk of malnutrition [18, 19]. Malnutrition remains a significant public health issue in Africa, with alarming statistics indicating that 38.6% of children under five years old are stunted, 28.4% are underweight, and 7.2% are wasted [20]. Globally, the prevalence rates of stunting, underweight, and wasting are estimated at 39.1%, 22.8%, and 4.1%, respectively [21]. Moreover, data from the Multiple Indicator Cluster Survey (MICS) in Sierra Leone revealed that 30% of children under the age of five were either underweight or severely underweight for their age, 40% were stunted, and 9% were wasted or too thin for their height [22]. These findings underscore the urgent need for targeted interventions to address malnutrition among vulnerable populations, particularly in regions affected by conflict and displacement. These children are particularly vulnerable to the effects of malnutrition, making them a key target for interventions aimed at improving overall nutritional status and health outcomes in these communities. The prevalence of malnutrition in young children can serve as an important indicator of the general population’s nutritional status, providing valuable data for health education and treatment initiatives. Addressing the underlying factors contributing to malnutrition, such as inadequate food aid and poor access to essential nutrients, is crucial for improving the health and well-being of refugees and displaced populations, particularly children who are most at risk.

Malnutrition in IDPs can lead to significant adverse effects on both their physical and mental health. Physically, malnutrition can result in nutritional deficiencies, gastrointestinal problems, weakness, and fatigue. Mentally, it can cause cognitive impairment, emotional distress, and social isolation Nutritional status serves as a critical public health indicator for refugees and displaced populations, as it is closely linked to the risk of short-term mortality. The complex causes of malnutrition in these vulnerable groups are influenced by a combination of dietary and environmental factors. Research focusing on IDPs has highlighted ongoing issues of under-nutrition, characterised by inadequate intake of both macro and micro-nutrients. While definitive causation has not been established, there is a strong likelihood that the insufficient food aid supplies provided during this period have contributed to the high rates of malnutrition, poor health outcomes, and increased mortality among displaced populations.

The study emphasises the urgent need for targeted health interventions for internally displaced persons in Sudan, highlighting the importance of addressing social determinants of health, providing gender-sensitive approaches, and improving access to mental health services. Recommendations include tailored interventions to mitigate anxiety and promote physical well-being among female IDPs, improving access to affordable and nutritious food and healthcare services for children, implementing psycho-social interventions, establishing mental health clinics in camps, increasing financing for healthcare programs, training more healthcare workers, improving accessibility to healthcare facilities, encouraging breastfeeding and maternal nutrition, and continuously monitoring mortality and malnutrition. Collaborative efforts from various stakeholders are essential in providing adequate care for IDPs. Prioritizing the physical health of IDPs is crucial for promoting their long-term health outcomes and successful integration into society. Addressing malnutrition comprehensively can enhance the resilience and recovery of displaced populations. Tailored interventions for diverse healthcare needs within different age groups and genders among displaced populations are necessary to address both mental and physical health needs, create employment opportunities, and improve the nutritional status of young children.

## LIMITATION

The limitations of the study include the inability to generalise findings in other regions of Sudan due to the cross-sectional design. The lack of evaluation of chronic diseases among internally displaced persons (IDPs) and difficulties in diagnosing physical health conditions in such situations also hindered the study. Additionally, there was a potential for respondents to exaggerate their suffering in hopes of receiving aid, even though it was made clear to the respondents before the interview that they would not receive any material benefits from the study and that the study team was not from an aid organization. Overall, the study emphasised the need for a more comprehensive approach to understanding health outcomes in conflict-affected populations.

## CONCLUSION

In conclusion, our study highlights the significant impact of internal displacement on the health of internally displaced people (IDPs) in Sudan, particularly in White Nile State. The high prevalence of mental illnesses, poor physical health, and malnutrition in IDP camps underscores the urgent need for comprehensive healthcare services, psycho-social support programs, and improvements to living conditions. Addressing these issues is crucial not only for the well-being of individual IDPs but also for public health in Sudan as a whole. Recommendations include increased funding for healthcare services, implementation of psycho-social support programs, improvements to living conditions in IDP camps, collaboration between government agencies and organizations, and redistributing displaced individuals to reduce crowding. Future research should continue to explore the relationships between displacement and health disparities through longitudinal studies, gender-specific studies, and resilience research to promote better health outcomes.

## Data Availability

All data produced in the present work are contained in the manuscript

## Acknowledgement

We are grateful to Alfia Nanda, for her assistance in reviewing the article. We are especially thankful to Altahir, Abobakar, Majda, and Hibat who assisted in data collection. We appreciate the support provided during the study by the White Nile State Ministry of Health. We thank the leaders of the IDP camps for helping the research team. We gratefully acknowledge the cooperation and involvement in the study from the IDP camp residents as well as the parents of the children. Lastly, we would like to express our gratitude to Kenana Sugar Company for providing funding for this research.

